# Denoised VEGFR2 expression relates to sunitinib efficacy in advanced Clear Cell Renal Cell Carcinoma

**DOI:** 10.1101/2021.11.10.21266155

**Authors:** Loïc Verlingue, Daphné Morel, Mickaël Schaeffer, Laurent Tanguy, Jordane Schmidt, Jean-Christophe Bernhard, Bertrand Loubaton, Dominique Bagnard

## Abstract

**Short summary:** Personalized biomarkers can facilitate decision making upon multiple therapeutic options in ccRCC. VEGFR2 expression denoised with 37 normal and tumor gene-expressions relates to sunitinib effect whereas raw VEGFR2 expression doesn’t relate to sunitinib effect.

**Background:** Several studies suggested that molecular analysis of patients with advanced clear cell renal cell carcinoma (ccRCC) could indicate whether a patient is susceptible of benefiting from sunitinib in first-line systemic treatment compared to immunotherapies. However, data remain conflicting and no predictive biomarker is validated so far to decipher if sunitinib could still represent a good therapeutic option in first line setting and beyond.

**Methods:** PREDMED^®^ denoised the tumor RNA expression of 37 genes including *KDR* (encoding VEGFR2) estimated by RT-qPCR, by normalizing it on the expression of normal kidney tissue and cell types. We investigated the performance of PREDMED^®^ VEGFR2-scoring to predict the clinical effect of sunitinib for patients affected by ccRCC.

**Results:** Among the 34 ccRCC patients’ samples retrospectively retrieved from the UroCCR project (NCT03293563), high VEGFR2 scores were associated with objective clinical responses under sunitinib treatment and low scores with stable disease or progression with a sensitivity of 86%, a specificity of 67% and an AUC of 72.5% (95%CI[50.1–94.9]; p=0.04). VEGFR2 scores were significantly and positively related to progression-free survival (HR = 0.465; 95%CI[0.221–0.978]; p=0.0311) and overall survival (HR = 0.400; 95%CI[0.192–0.834]; p=0.0134) under sunitinib treatment. In our cohort, raw VEGFR2 expression (before PREDMED^®^ processing) was not related to the above mentioned outcomes.

**Conclusion:** We describe a gene-expression based algorithm that is accurately related to the effect of sunitinib for patients with ccRCC. We further plan a validation of PREDMED^®^ for combinatorial strategies involving antiangiogenics and immune-checkpoint blockers.

## 1. INTRODUCTION

Renal cell carcinoma (RCC) corresponds to 85% of all kidney cancer, with clear cell renal cell carcinoma (ccRCC) being the most frequent subtype accounting up to 80% of all RCC ^1^. The molecular characterization of sporadic ccRCC is highly specific, with the *Von Hippel-Lindau* (*VHL*) gene being altered or epigenetically silenced in more than 90% of the cases ^2,3^. The loss of VHL leads to the stabilization of hypoxia inducible factors (HIF-1α and HIF-2α), stimulating the production of oncogenic and pro-angiogenic agents such as VEGF and PDGF ^4,5^ that drive the majority of ccRCC and is efficiently targeted using antiangiogenics.

CcRCC often remains asymptomatic for several years and more than half of ccRCC are diagnosed incidentally ^6^, typically at an advanced stage. The management of advanced and metastatic ccRCC mostly relies on systemic treatments according to a risk stratification that split patients into good-, intermediate- and poor-prognosis groups following International Metastatic RCC Database Consortium (IMDC criteria) ^7,8^. In the past few months, clinical practice guidelines drastically evolved to propose as the preferred first-line regimen PD-1 inhibitor (pembrolizumab or nivolumab) for all risk-groups patients, combined or not with an antiangiogenic (axitinib) or CTLA-4 inhibitor (ipilimumab). Though, sunitinib and pazopanib – both antiangiogenic multikinases – still represent recommanded therapeutic alternative options for first-line systemic treatment, for example for patients ineligible or unwiling to receive immune-checkpoint blockers (NCCN Guidelines for Kidney Cancer, version 1.2021 – July 15, 2020).

Some pivotal trials demonstrated the clinical superiority of immune-checkpoint inhibitors used in combination over monotherapies of anti-angiogenics in first-line setting in unselected patients ^9,10^. However, several studies suggested that some patients might benefit more from a monotherapy of antiangiogenic in first line than from immune-checkpoint blockers, combined or not with antiangiogenics, based on their tumoral molecular profiling. For example, *Liu et al*. reported that PBRM1 loss-of-function – which is found in 40% of ccRCC ^3^ – was associated with an upregulated angiogenesis and a less immunogenic microenvironement, and therefore patients with *PBRM1*-mutated ccRCC were more likely to benefit from first-line sunitinib than an immune-checkpoint blocker ^11^. These findings were consistent with the results of the prospective IMmotion150 trial that reported improved survival outcomes following sunitinib compared to atezolizumab (anti-PDL1) with or without bevacizumab in molecularly selected patients, based on a gene expression signature of 7 VEGF-inducible angiogenesis-associated genes ^12^. However, the correlation between expression of VEGF or VEGF-related proteins and response to sunitinib remains unclear and conflicting data limit its application in the clinic ^13–16 17,18^.

In this study, we explored the clinical performance of a method to denoise the analysis of VEGFR2 expression in ccRCC Formalin-Fixed Paraffin-Embedded (FFPE) samples, based on a 37 gene-expression signature from the tumor and normal kidney cells. We evaluated if VEGFR2 scores could predict objective responses and outcomes of patients with advanced or metastatic ccRCC treated with sunitinib in first-line, second-line or third-line.

## 2. MATERIALS AND METHODS

### 2.1. Data collection and patient selection

Patients were retrospectively retrieved from the UroCCR project (French research network on kidney Cancer - NCT03293563). Eligible patients were 18 years of age or older, had a primary or recurrent ccRCC treated with sunitinib in any treatment line setting, with available material from surgical resection of the primary tumor prior to sunitinib treatment. Patients were excluded from the analysis in case of missing clinical data or RNA Integrity Number (RIN) below 7.

Outcomes collected were the RECIST V1.1 best response: complete or partial response (CR or PR, respectively), stable disease (SD) or progressive disease (PD); PFS in months (calculated as the time from the first intake of sunitinib to the date of first documented progression or death); OS in months (calculated as the time from the first intake of sunitinib to the date of death from any cause); duration of follow-up; number and type of previous treatment lines. Objective response rate (ORR) was defined as the proportion of patients with CR or PR under sunitinib ^19^. Additional clinical characteristics available comprised: age at diagnosis, gender, African phenotype ethnicity (yes/no), number and type of previous lines of treatment and the type of surgery (cytoreductive of complete nephrectomy).

### 2.2. Study objectives

The primary objective was to evaluate the relation between PREDMED® VEGFR2-scores and the effect of sunitinib, represented by ORR and PFS, for patients with advanced or metastatic ccRCC. The secondary objective was to evaluate the relation between PREDMED® VEGFR2-scores and OS. Performance estimation relied on sensitivity, specificity and Receiver Operating Characteristic Area Under the Curve (AUC), as specified by the STARD 2015 guidelines ^20^.

### 2.3. PREDMED Medical Device

#### 2.3.1. Rationale

PREDMED® is an *in vitro* diagnostic multivariate index assay (IVDMIA) that normalizes the RNA expression of 37 selected genes from tumor samples on a bank of normal tissues and cells’ gene expressions. The algortihm provides a score for each gene, ranked from 0 to 1000, that reflects their relative dysregulations compared to normal values, as previously described ^21^.

The provisional limited panel of 37 genes was selected based on current knowledge of mainly targetable biological mechanisms implicated in ccRCC, comprising the tumor cells, stromal cells, vessels and immune cells. (listed in **Fig. 1**). For the current study we prospectively choosed to use the VEGFR2-score only.

**Figure 1:**
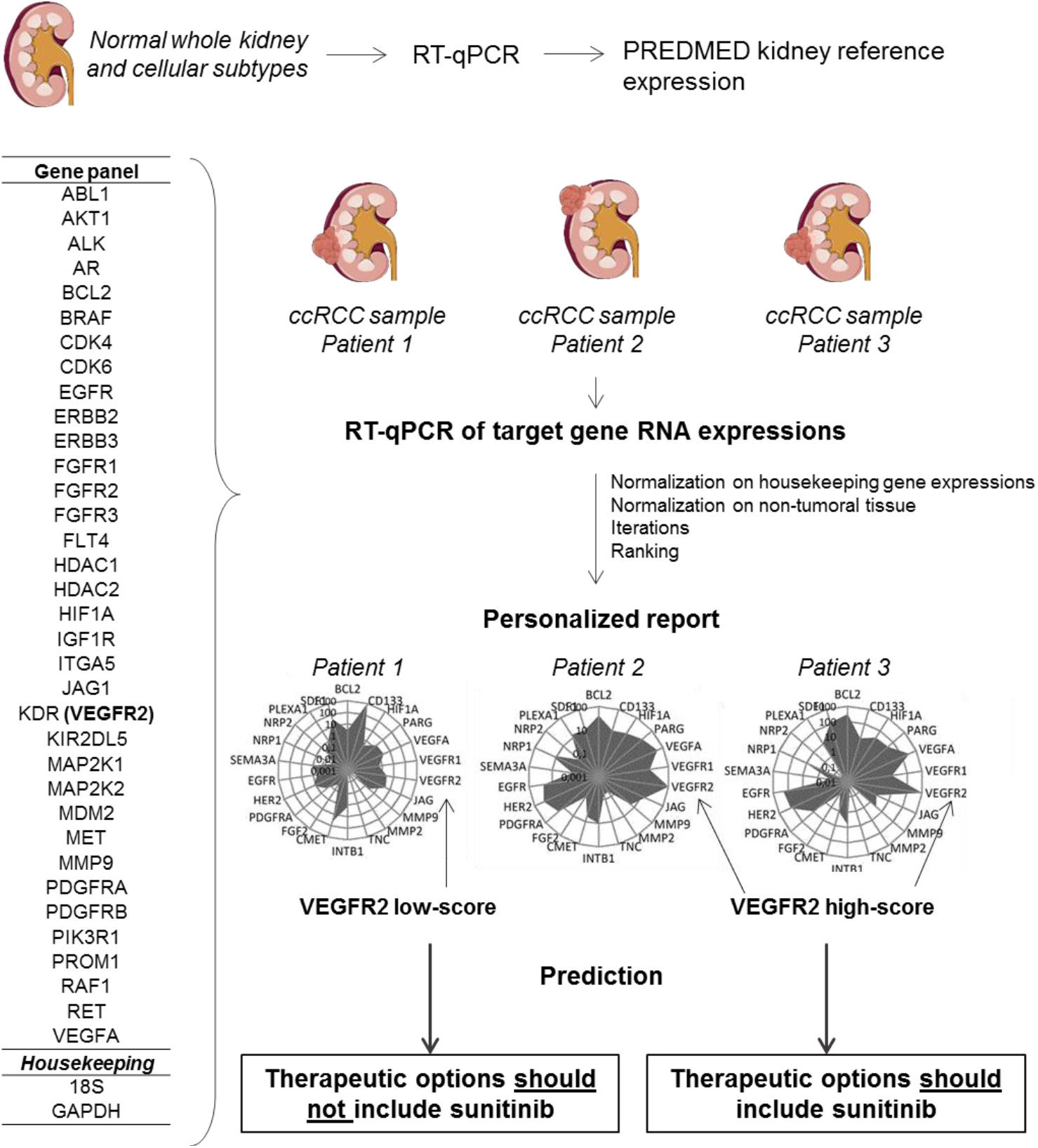
Summary of PREDMED method. PREDMED normalizes the RNA expression of 37 genes from tumor samples on a bank of normal tissues and cells’ gene expressions. The algortihm provides a score for each gene, ranked from 0 to 1000, that reflects their relative dysregulations compared to normal values.

#### 2.3.2. Samples and biological methods

CcRCC FFPE samples (CRB-K - CHU Bordeaux) were all processed in a centralized laboratory in Strasbourg, France (INSERM U1119, BMNST Lab, University of Strasbourg, Labex Medalis, Fédération de Médecine Translationnelle). Blocks were stored at -20°C and RNA samples were stored at -80°C following extraction. Total RNA was extracted with TRI Reagent^®^ solution (Molecular Research Center; #TR118), quantified, assessed for quality (RIN) and reverse transcribed (Applied Biosystems; #4368814). The obtained cDNA was diluted to get a final concentration of 1 μg /100 μL. RT-qPCR was performed using TaqMan Gene expression Master Mix (Applied Biosystems; #4369016). Experiments were conducted using customized microplates specially designed for this project by Applied Biosystems (Custom TaqMan Array Plates; #4391526) to contain human specific TaqMan^®^ probes at 1X and primers at 1X (list provided in **Suppl Fig. 1**).

#### 2.3.3. Analytical methods / PREDMED® algorithm

PREDMED® normalization method has been previously described ^21^. Briefly, the reference panel gene expressions were assessed using a cocktail of non-tumoral kidney tissues which comprised: whole normal kidney total RNA (#AM7976), medullary kidney RNA (CRB-K - CHU Bordeaux), cortical kidney RNA (CRB-K - CHU Bordeaux), human renal glomerular endothelial cell total RNA (#4005-SC), human renal proximal tubular epithelial cell total RNA (#4105-SC), human renal cortical epithelial cell total RNA (#4115-SC), human renal epithelial cell total RNA (# 4125-SC), human renal mesangial cell total RNA (#4205-SC), and a low grade carcinoma of kidney total RNA (# CR559126).

Then, to reduce interindividual variability and allow the normalization process on the reference panel, gene expressions were first normalized on the mean expression of two housekeeping genes (*18S ribosomal RNA* and *Glyceraldehyde 3-phosphate dehydrogenase GAPDH*), as follow: ΔCt (gene) = Ct (gene) – mean Ct (housekeeping genes). 2^-ΔCt (gene)^.

Following similar normalization steps, run multiple times, the resulting score ranged from 0 to 1000 for each gene: 1000 corresponding to the highest relative upregulation from normal. PREDMED® is protected by International Application patent PCT/EP2016/078353.

### 2.4. Statistical analysis

All recorded variables were described by using position and dispersion statistics, such as mean, median and 95% confidence interval (95%CI). The assumption of normality (defined by the Gaussian distribution) was tested by the Shapiro Wilk test on each quantitative variable distribution. To compare survival distributions, we used the log-rank Mantel-Cox test, and described the results with the Hazard Ratio, 95%CI ratio and associated p-value. All statistical tests were two-tailed and a p-value < 0.05 was considered as statistically significant. All analyses were performed using R software under its version 3.1 and JAGS for the MCMc estimations in Bayesian models ^22^. Except for ROC curves generated using R, all graphs were created using GraphPad Prism® V8.0.2.

### 2.5. Ethics

The trial was conducted in accordance with the local Good Clinical Practice guidelines (CNIL number declaration 2005853 v 0, DC-2017-3040). The biobank biological resource center number associated with this study is BB-0033-00036. The UroCCR project (NCT03293563) obtained the authorization number DR-2013-206 from the national information science and liberties commission (CNIL) and all patients included consented to the use of their personal and genetic data.

## 3. RESULTS

### 3.1. Patients’ characteristics

We retrospectively collected 46 FFPE tumor samples from patients addressed for advanced or metastatic ccRCC between December, 2006 and February, 2016 (**Fig. 1**). Among these 46 patients, 5 patients were excluded from the analysis: 2 patients received sunitinib before surgical resection, 1 patient never received sunitinib and 2 patients had missing clinical data. Gene expressions were assessed by RT-qPCR and led to the exclusion of 7 patients due to poor RNA quality. Altogether, 34 patients remained (**Fig. 2)**. The median age was 66 years old and the sex ratio approximately 3 males for 1 woman (**Table 1**). The majority of patients (N=30, 88.2%) had a ccRCC at a metastatic stage at the time of study and 31 (91.2%) patients received sunitinib as first-line systemic treatment.

**Table 1:**
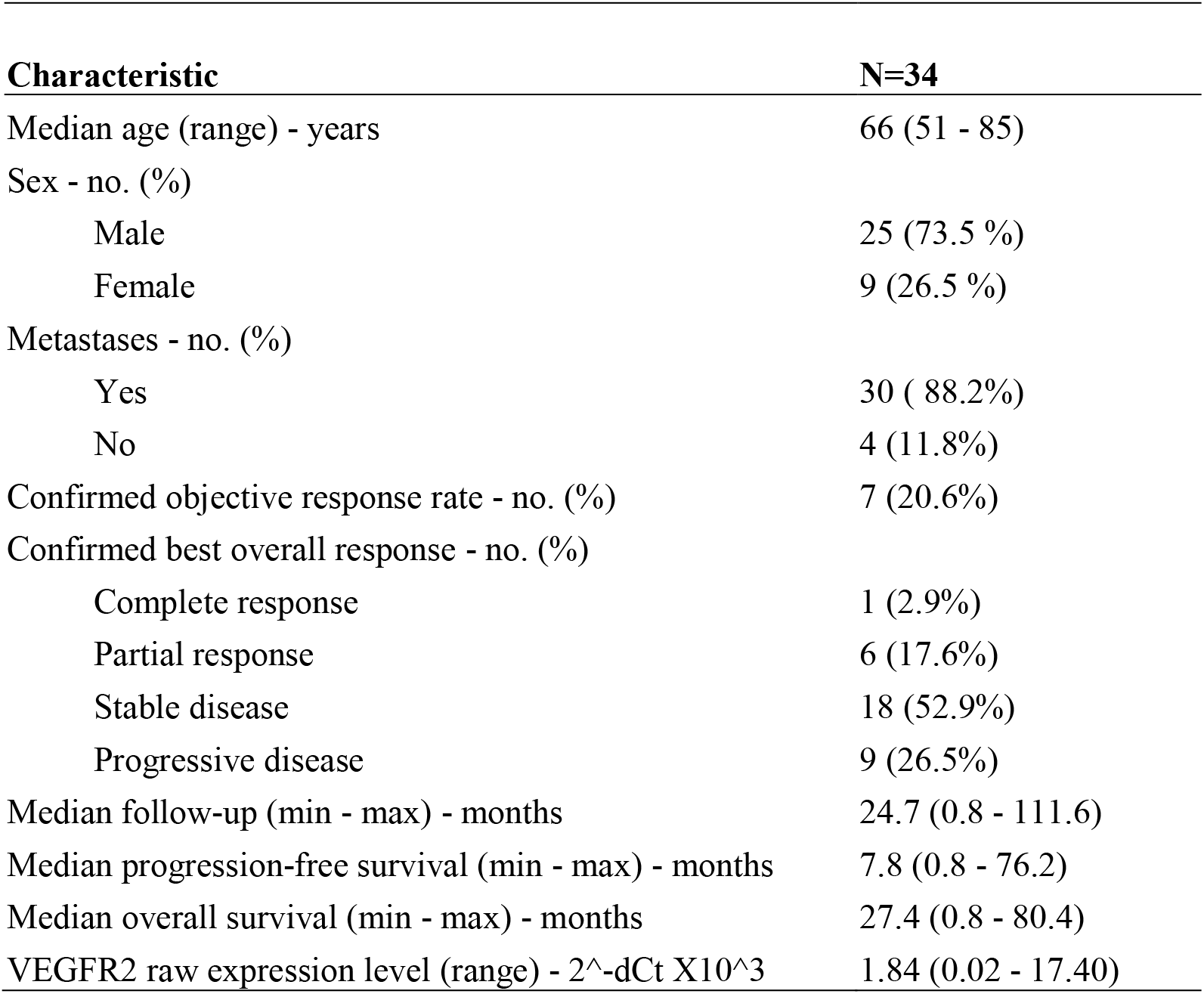
Baseline patient characteristics and clinical outcomes of all patients treated with sunitinib and included in the gene expression analysis (N=34)

**Figure 2:**
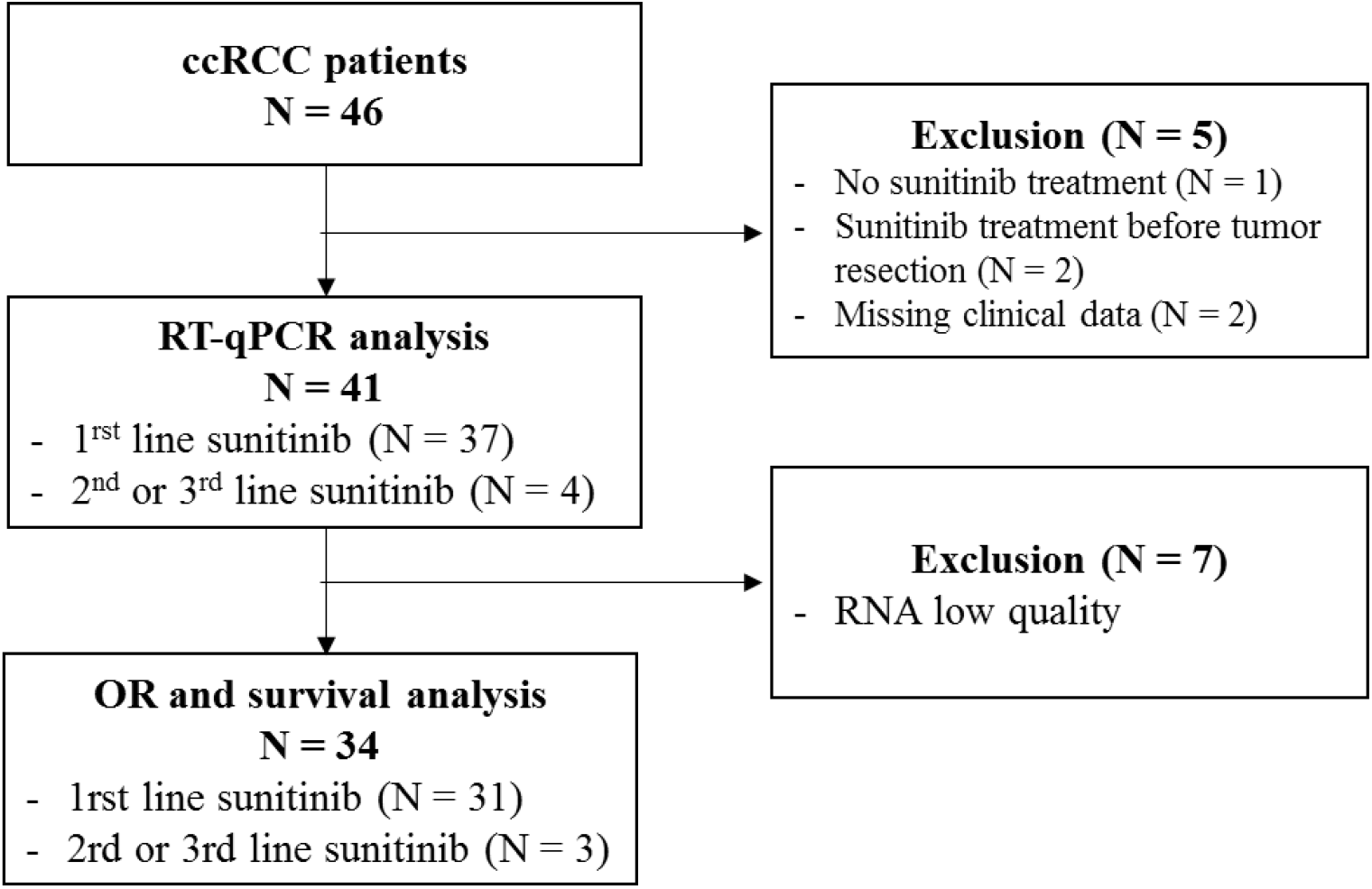
Flowchart of the study.

In our cohort, sunitinib treatment was associated with an overall objective response rate of 20.6%, including 1 complete response (2.9%) and 6 partial responses (17.6%). Nine patients (26.5%) progressed and 18 patients (52.9%) harbored stable disease as best response. After a median follow-up of 24.7 months, median PFS was 7.8 months and median OS was 20.2 months, which is similar to data from pivotal trials that evaluated sunitinib in the first-line setting ^9,10^. At the time of end of study, 5 patients were still alive, 2 of them still under sunitinib treatment.

### 3.2. VEGFR2-score and response to sunitinib

VEGFR2-scores ranged between 2.0 and 1000.0, with a mean of 504.3. Six out of 7 patients with PR or CR had high VEGFR2-score, and 18 out of 27 patients with PD or SD had low VEGFR2-score. It resulted in a sensitivity of 86%, a specificity of 67%, an AUC of 72.5% (95%CI [50.1 – 94.9]; p=0.04) (**Fig. 3A,B**). On the ten patients with the highest VEGFR2-scores, 1 had a complete response, 3 had partial responses and 6 had stable diseases. Conversely, low VEGFR2-scores had a negative predictive value of 94.7%. Raw VEGFR2 expression had poorer relation to response to sunitinib compared to PREDMED® VEGFR2-scores, with an AUC of 48.4% (95%CI [25.2 – 71.6]; p=0.32), a sensitivity of 71% and a specificity of 56% (**Fig. 3C,D**).

**Figure 3:**
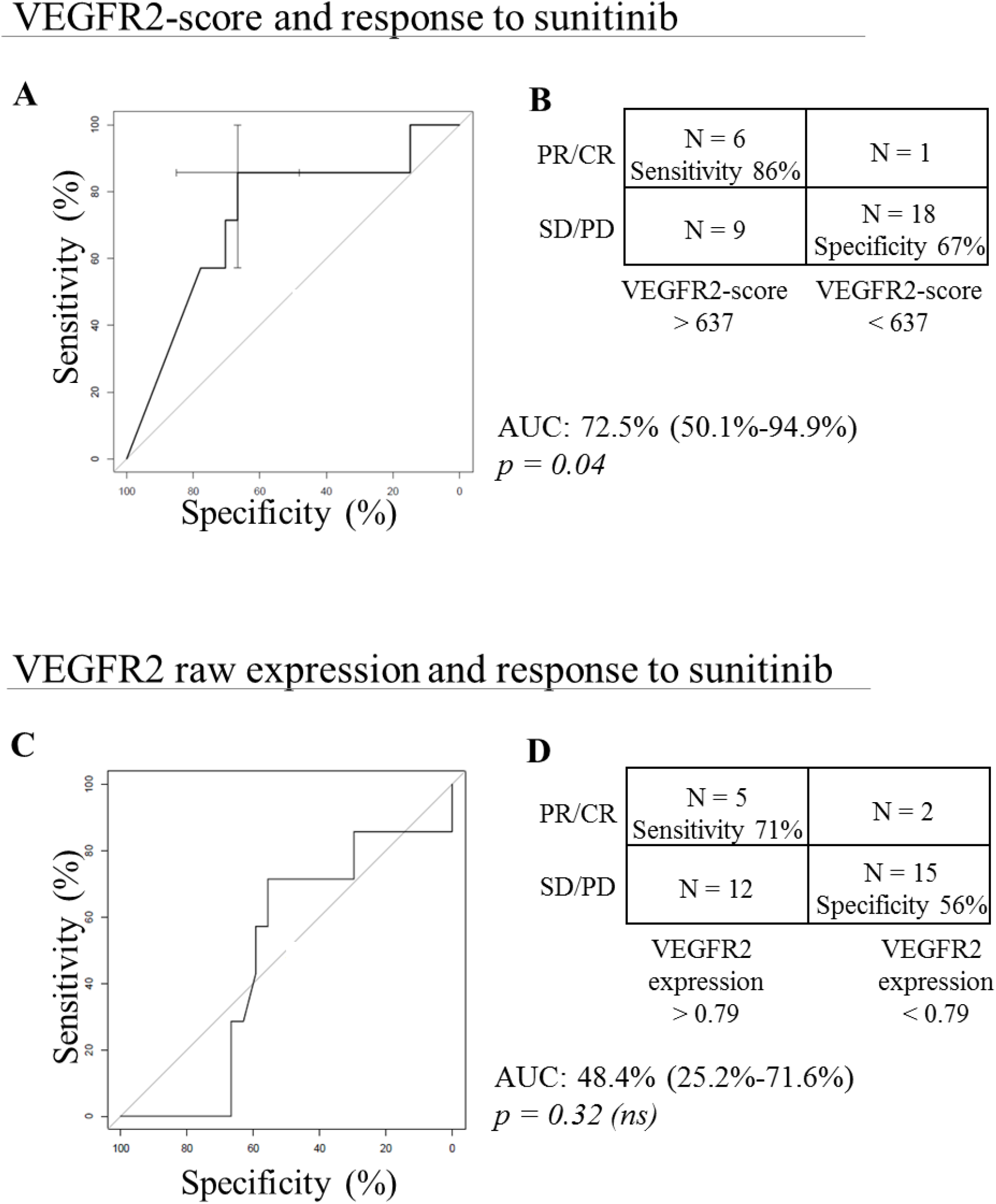
VEGFR2 and response to sunitinib. **A**, VEGFR2-score and response to sunitinib: ROC curve displaying the prediction performances of the VEGFR2-score computed using the PREDMED® signature algorithm. Area under the curve (AUC), 95% confidence interval and associated p-value are indicated. **B**, Contingency table depicting the number of partial and complete responses (PR/CR) and stable and progressive diseases (SD/PD) accurately predicted using the VEGFR2-score with a cut-off at 637. **C**, VEGFR2 raw expression and response to sunitinib: ROC curve displaying the prediction performances of the VEGFR2 mRNA expression alone after normalization on housekeeping genes. Area under the curve (AUC), 95% confidence interval and associated p-value are indicated. **D**, Contingency table depicting the number of partial and complete responses (PR/CR) and stable and progressive diseases (SD/PD) accurately predicted using the VEGFR2 mRNA expression with a cut-off at 0.79.

### 3.3. VEGFR2-scores and outcome under sunitinib

We observed that PFS was significantly longer in patients with higher VEGFR2-scores (HR: 0.465, 95%CI [0.221–0.978], p=0.0311) (**Fig. 4A**). OS was also significantly longer in patients with higher VEGFR2-scores (HR: 0.400, 95%CI [0.192–0.834], p=0.0134) (**Fig. 4B**). The 5 patients who were still alive at the date of end of study – more than 77.2 months after the initiation of sunitinib – had very high VEGFR2-score (4 with 1000, 1 with 749). Among them, 2 patients with higest scores were still under sunitinib treatment, 81.4 and 92.9 months after initiation. Conversely, raw VEGFR2 mRNA expression was not significantly related to PFS and OS (for PFS, HR = 0.682, 95%CI [0.323-1.438], p=0.28; and for OS, HR = 0.705, 95%CI [0.338-1.470], p=0.33) (Supplementary Figure 2).

**Figure 4:**
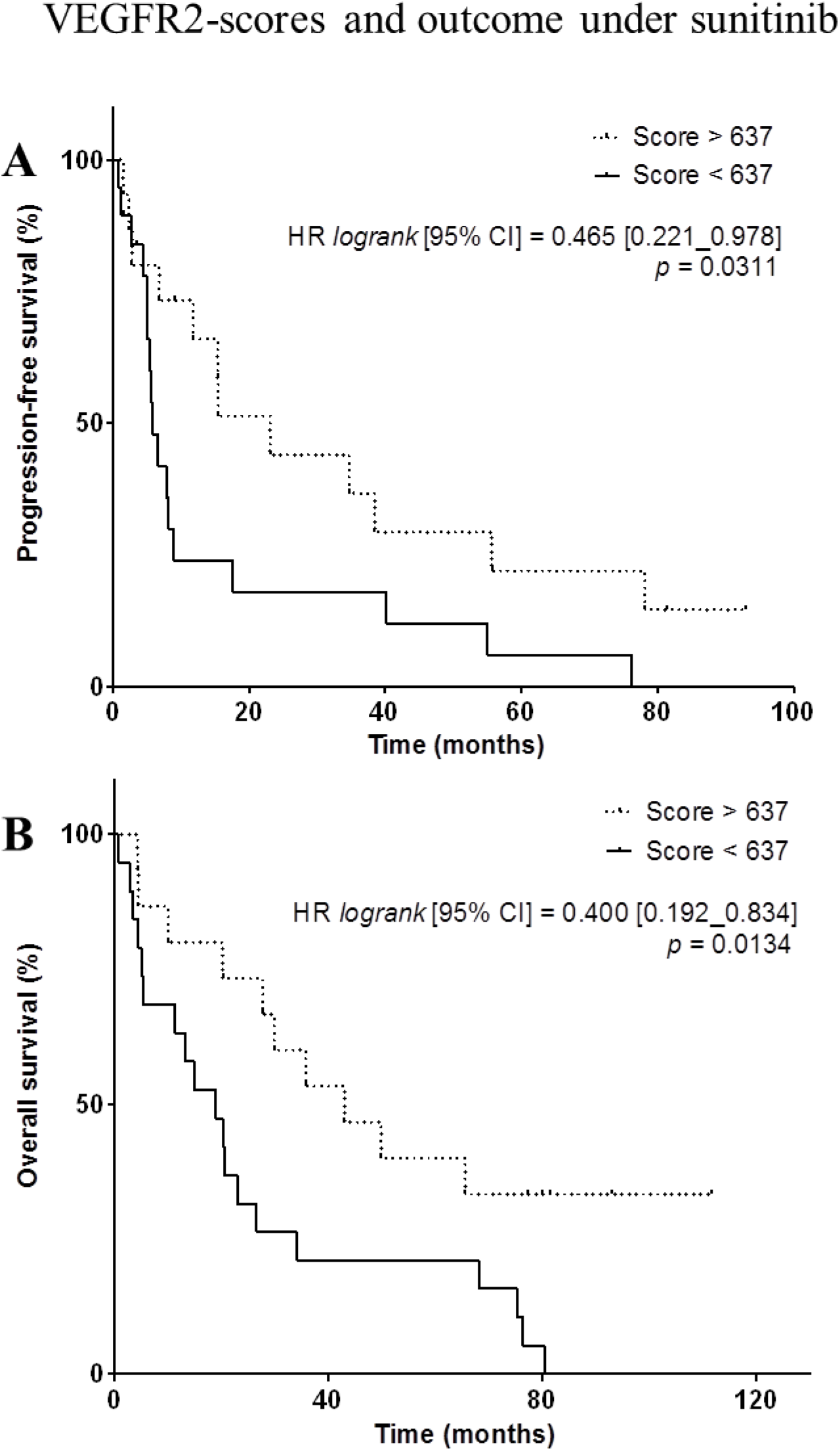
VEGFR2-scores and outcome under sunitinib. **A**,**B**, Kaplan-Meier analysis of progression-free survival (**A**) and overall survival (**B**) of patients with a high VEGFR2-score (> 637, dotted line) *versus* patients with a low VEGFR-score (< 637, full line). Hazard ratio (logrank), 95% confidence interval and associated p-value are indicated.

## 4. DISCUSSION

PREDMED® denoised the tumor expression of VEGFR2 by analyzing the tumor and normal kidney tissues and cell types of 37 gene expressions selected for their biological and therapeutic roles. For 34 patients with advanced of metastatic ccRCC who received sunitinib, VEGFR2-scores related to overall response rate, progression-free survival and overall survival. Six out of 7 patients who responded to sunitinib had a high VEGFR2-score (sensitivity 86%), 18 out of 27 patients with stable or progressive disease displayed a low VEGFR2-score (specificity 67%). In addition, wrong negative prediction only occurred with one patient who showed partial response while displaying a low VEGFR2-score (negative predictive value 94.7%). It included patient in various treatment lines with sunitinib, independently from the prognosis-risk groups. Previous studies suggested that high VEGFR2 expression may reflect favorable outcome on sunitinib in patients with ccRCC, and therefore could be used as a predictive biomarker of response ^13,14,16,17^. In our cohort, the raw VEGFR2 expression assessed by RT-qPCR failed to relate to the above mentioned outcomes.

Our study suggest that conventional gene expression analysis to drive targeted treatment relies may be limited by inherent noise. Noise may come from biological sampling, inter-individual variation, or technical variation, among other factors. Denoising expression data from internal and/or external factors is not a usual approach. One of the few clinical evaluation of such hypothesis has been performed in the WINTHER study ^23^ (NCT01856296). WINTHER proposed transcriptomic analysis from tumor biospsies which were normalized on normal surounding tissue of various cancer types. Although the study did not meet its pre-specified primary end-point, it yielded promising outcome results in heavily pre-treated patients and confirmed that assessing the expression profile of tumor to guide treatment is feasible and do not delay therapeutic care. Our tool differs from the WINTHER algorithm through its iterative multi-normalization process and a large number of reference normal tissue gene expressions. It does not require the biopsy of healthy tissue from the patients to compute the score.

The present study is limited by its retrospective nature; thus, a prospective validation of our findings is planned through a non-interventional study on advanced ccRCC patients receiving sunitinib,. We also consider evaluating this strategy for other ccRCC-approved antiangiogenics, in particular to define whether another antiangiogenic, such as pazopanib or axitinib, should be preferred over sunitinib or should be avoid as well in case of low VEGFR2-scoring. Another limitation of the study is the scarce clinical characteristics available in the database. We could not calculate the standard prognosis IMDC scores to investigate PREDMED® predictions within each risk-group of patients. Nevertheless, our approach shows that gene expression assessment from surgical samples can relate to outcome under sunitinib treatment when sophisticated normalization is performed. Finally, in the current study, we did not take into consideration the scores associated with other genes targeted by sunitinib, such as PDGFR or RET. Future multivariate development of the algorithm could allow more specific multi-kinase predictions.

Given the gene panel used and its potential versatility, PREDMED® test can address various therapeutic options, including targeted therapies and immunotherapies, in various tumor types. In this pilot study, the highest priority was given to advanced and metastatic ccRCC, as it remains one of the tumor types with the largest approved treatment options with no validated biomarker available. An additional attractive perspective concerns combinatorial strategies involving immune-checkpoint blockers with or without antiangiogenics in first-line setting, particularly in intermediate and high-risk patients. It is conceivable that a small – and yet unidentified – proportion of patients may benefit from an antiangiogenic added to the anti-PD(L)1 drug, and conversely, some patients may more benefit from a doublet of immune-checkpoint blockers. Importantly, some patients may also benefit from a monotherapy of antiangiogenic and be primarily resistant to immune-checkpoint blockers ^11,12^. Biomarkers are urgently needed to identify such a population; hence, to ease personalized decision making and to optimize therapeutic care for advanced and metastatic cancer patients.

## Data Availability

All data produced in the present study are available upon reasonable request to the authors

## ACKNOWLEDGMENTS

The authors thank the members of the UroCCR biological resources center (CRB-K) at Bordeaux, France. The authors are gratefull to Laurence Albiges for her advices and support for this work. The authors would like to thank Justine Fritz, Mathilde Baranger and Coralie Gianesini for their contribution to the data generation.

## CONFLICT OF INTEREST

LV, DM, MS, LT, BL and DB report personal fees from Adaptherapy related to the submitted work.

LV reports grants from Bristol-Myers Squibb, non-personal fees from Servier and Pierre-Fabre, outside of the submitted work.

LV, as part of the Drug Development Department (DITEP):

Principal/sub-Investigator of Clinical Trials for Abbvie, Adaptimmune, Aduro Biotech, Agios Pharmaceuticals, Amgen, Argen-X Bvba, Arno Therapeutics, Astex Pharmaceuticals, Astra Zeneca Ab, Aveo, Basilea Pharmaceutica International Ltd, Bayer Healthcare Ag, Bbb Technologies Bv, Beigene, Blueprint Medicines, Boehringer Ingelheim, Boston Pharmaceuticals, Bristol Myers Squibb, Ca, Celgene Corporation, Chugai Pharmaceutical Co, Clovis Oncology, Cullinan-Apollo, Daiichi Sankyo, Debiopharm, Eisai, Eisai Limited, Eli Lilly, Exelixis, Faron Pharmaceuticals Ltd, Forma Tharapeutics, Gamamabs, Genentech, Glaxosmithkline, H3 Biomedicine, Hoffmann La Roche Ag, Imcheck Therapeutics, Innate Pharma, Institut De Recherche Pierre Fabre, Iris Servier, Janssen Cilag, Janssen Research Foundation, Kura Oncology, Kyowa Kirin Pharm. Dev, Lilly France, Loxo Oncology, Lytix Biopharma As, Medimmune, Menarini Ricerche, Merck Sharp & Dohme Chibret, Merrimack Pharmaceuticals, Merus, Millennium Pharmaceuticals, Molecular Partners Ag, Nanobiotix, Nektar Therapeutics, Novartis Pharma, Octimet Oncology Nv, Oncoethix, Oncopeptides, Orion Pharma, Ose Pharma, Pfizer, Pharma Mar, Pierre Fabre, Medicament, Roche, Sanofi Aventis, Seattle Genetics, Sotio A.S, Syros Pharmaceuticals, Taiho Pharma, Tesaro, Xencor. Research Grants from Astrazeneca, BMS, Boehringer Ingelheim, Janssen Cilag, Merck, Novartis, Onxeo, Pfizer, Roche, Sanofi. Non-financial support (drug supplied) from Astrazeneca, Bayer, BMS, Boringher Ingelheim, Medimmune, Merck, NH TherAGuiX, Onxeo, Pfizer, Roche.

The other authors have no conflict of interest to declare.

## SUPPLEMENTARY FIGURES

**Supplementary figure 1:**
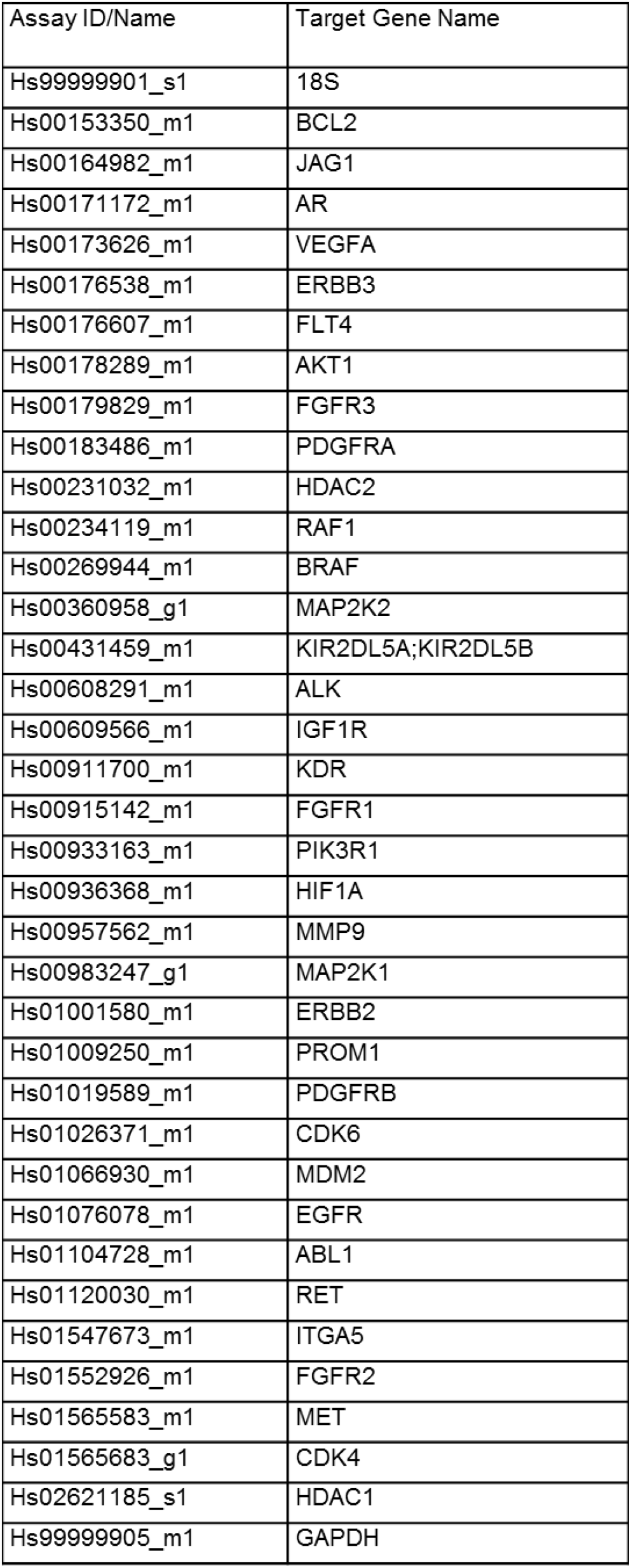
List of the targeted genes evaluated by RT-qPCR to enrich the algorithm and corresponding primer

**IDs.Supplementary figure 2:**
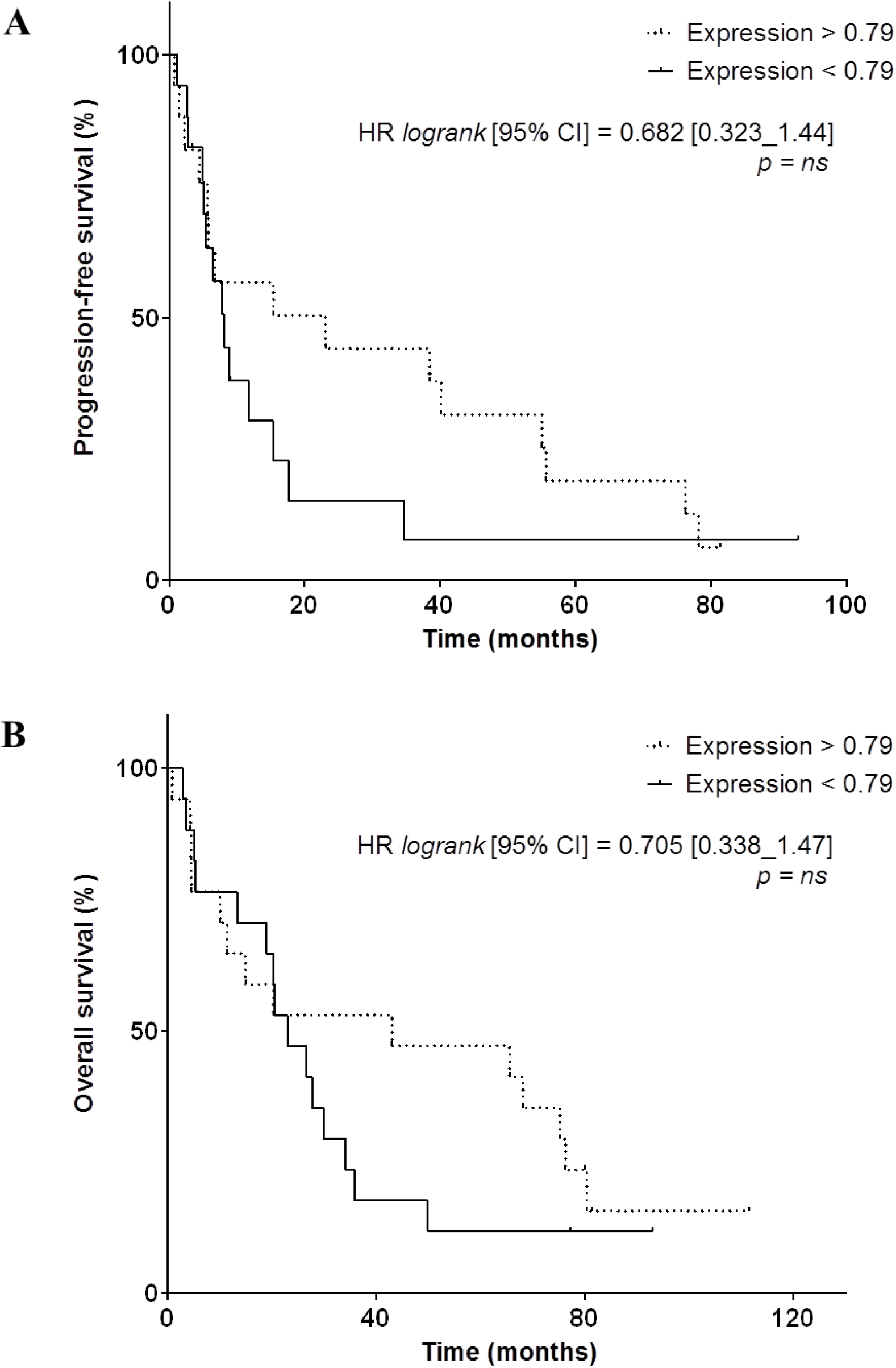
**A**,**B**, Kaplan-Meier analysis of progression-free survival (**A**) and overall survival (**B**) of patients displaying a high VEGFR2 mRNA expression (> 0.79, dotted line) *versus* patients displaying a low VEGFR2 mRNA expression (< 0.79, full line). Hazard ratio (logrank), 95% confidence interval and associated p-value are indicated.

